# Artificial intelligence driven assessment of routinely collected healthcare data is an effective screening test for COVID-19 in patients presenting to hospital

**DOI:** 10.1101/2020.07.07.20148361

**Authors:** Andrew AS Soltan, Samaneh Kouchaki, Tingting Zhu, Dani Kiyasseh, Thomas Taylor, Zaamin B. Hussain, Tim Peto, Andrew J Brent, David W. Eyre, David Clifton

**Affiliations:** John Radcliffe Hospital, Oxford University Hospitals NHS Foundation Trust; Oxford University Clinical Academic Graduate School, University of Oxford; Institute of Biomedical Engineering, Dept. Engineering Science, University of Oxford; Harvard Graduate School of Education, Harvard University; Harvard T.H. Chan School of Public Health, Harvard University; Centre for Vision, Speech and Signal Processing, University of Surrey; Nuffield Department of Medicine, University of Oxford; Big Data Institute, Nuffield Department of Population Health, University of Oxford

**Keywords:** SARS-CoV-2, COVID-19, Artificial Intelligence, Machine Learning, Screening test, Diagnosis, Electronic Health Records, Emergency Department

## Abstract

**Background:** Rapid identification of COVID-19 is important for delivering care expediently and maintaining infection control. The early clinical course of SARS-CoV-2 infection can be difficult to distinguish from other undifferentiated medical presentations to hospital, however for operational reasons SARS-CoV-2 PCR testing can take up to 48 hours. Artificial Intelligence (AI) methods, trained using routinely collected clinical data, may allow front-door screening for COVID-19 within the first hour of presentation.

**Methods:** Demographic, routine and prior clinical data were extracted for 170,510 sequential presentations to emergency and acute medical departments at a large UK teaching hospital group. We applied multivariate logistic regression, random forests and extreme gradient boosted trees to distinguish emergency department (ED) presentations and admissions due to COVID-19 from pre-pandemic controls. We performed stepwise addition of clinical feature sets and assessed performance using stratified 10-fold cross validation. Models were calibrated during training to achieve sensitivities of 70, 80 and 90% for identifying patients with COVID-19. To simulate real-world performance at different stages of an epidemic, we generated test sets with varying prevalences of COVID-19 and assessed predictive values. We prospectively validated our models for all patients presenting or admitted to our hospital group between 20^th^ April and 6^th^ May 2020, comparing model predictions to PCR test results.

**Results:** Presentation laboratory blood tests, point of care blood gas, and vital signs measurements for 115,394 emergency presentations and 72,310 admissions were analysed. Presentation laboratory tests and vital signs were most predictive of COVID-19 (maximum area under ROC curve [AUROC] 0.904 and 0.823, respectively). Sequential addition of informative variables improved model performance to AUROC 0.942.

We developed two early-detection models to identify COVID-19, achieving sensitivities and specificities of 77.4% and 95.7% for our ED model amongst patients attending hospital, and 77.4% and 94.8% for our Admissions model amongst patients being admitted. Both models offer high negative predictive values (>99%) across a range of prevalences (<5%). In a two-week prospective validation period, our ED and Admissions models demonstrated 92.3% and 92.5% accuracy (AUROC 0.881 and 0.871 respectively) for all patients presenting or admitted to a large UK teaching hospital group. A sensitivity analysis to account for uncertainty in negative PCR results improves apparent accuracy (95.1% and 94.1%) and NPV (99.0% and 98.5%). Three laboratory blood markers, Eosinophils, Basophils, and C-Reactive Protein, alongside Calcium measured on blood-gas, and presentation Oxygen requirement were the most informative variables in our models.

**Conclusion:** Artificial intelligence techniques perform effectively as a screening test for COVID-19 in emergency departments and hospital admission units. Our models support rapid exclusion of the illness using routinely collected and readily available clinical measurements, guiding streaming of patients during the early phase of admission.

**Brief:** The early clinical course of SARS-CoV-2 infection can be difficult to distinguish from other undifferentiated medical presentations to hospital, however viral specific real-time polymerase chain reaction (RT-PCR) testing has limited sensitivity and can take up to 48 hours for operational reasons. In this study, we develop two early-detection models to identify COVID-19 using routinely collected data typically available within one hour (laboratory tests, blood gas and vital signs) during 115,394 emergency presentations and 72,310 admissions to hospital. Our emergency department (ED) model achieved 77.4% sensitivity and 95.7% specificity (AUROC 0.939) for COVID-19 amongst all patients attending hospital, and Admissions model achieved 77.4% sensitivity and 94.8% specificity (AUROC 0.940) for the subset admitted to hospital. Both models achieve high negative predictive values (>99%) across a range of prevalences (<5%), facilitating rapid exclusion during triage to guide infection control. We prospectively validated our models across all patients presenting and admitted to a large UK teaching hospital group in a two-week test period, achieving 92.3% (n= 3,326, NPV: 97.6%, AUROC: 0.881) and 92.5% accuracy (n=1,715, NPV: 97.7%, AUROC: 0.871) in comparison to RT-PCR results. Sensitivity analyses to account for uncertainty in negative PCR results improves apparent accuracy (95.1% and 94.1%) and NPV (99.0% and 98.5%). Our artificial intelligence models perform effectively as a screening test for COVID-19 in emergency departments and hospital admission units, offering high impact in settings where rapid testing is unavailable.

## Background

Severe Acute Respiratory Syndrome Coronavirus 2 (SARS-CoV-2), a novel coronavirus, is responsible for the Coronavirus Disease-2019 (COVID-19) pandemic of 2020^1^. The early clinical course of COVID-19, which often includes common symptoms such as fever and cough, can be challenging for clinicians to distinguish from other respiratory illnesses ^2–4^.

Testing for SARS-CoV-2 through real-time polymerase chain reaction (RT-PCR) assay of nasopharyngeal swabs, most commonly targeting the viral RNA-dependent, RNA polymerase (RdRp) or nucelocapsid genes, has been widely adopted, but has limitations ^3,5,6^. These include limited sensitivity^5,7^, prolonged turnaround time of up to 72 hours in some centres, and requirements for specialist laboratory infrastructure and expertise^8^. There therefore exists an urgent clinical need for rapid, point-of-care identification of COVID-19 to support expedient delivery of care, and assist front door triage and patient streaming for infection control purposes^9^.

The increasing use of electronic healthcare record (EHR) systems in hospitals has improved the richness of available clinical datasets available to study COVID-19. However, many studies to date have relied on manual collection of selected clinical variables^10–12^. In contrast, high-throughput electronic data extraction and processing techniques can enable curation of rich datasets from EHRs^13^, incorporating all clinical data available on presentation, and may combine with advanced machine learning techniques to produce a rapid screening tool for COVID-19 that fits within existing clinical care pathways^11,14^.

Approaches to produce a rapid screening tool, with utility during the early phase of hospital presentations, should use only clinical data available prior to the point of prediction^15^. Basic laboratory blood test data and physiological clinical measurements (vital signs) are amongst routinely collected healthcare data typically available within the first hour of presentation to hospital, and patterns of changes have been described in retrospective, observational studies of COVID-19 patients (variables including lymphocyte count, ALT, CRP, D-Dimer and Bilirubin^3,4,16,17^). Moreover, prior healthcare data available within the EHR may have utility in identifying risk factors for COVID-19 or underlying conditions which may cause alternative, but similar presentations.

We applied artificial intelligence methods to a rich clinical dataset with the aim of developing a rapidly deployable model for identifying and ruling out COVID-19 using routinely collected healthcare data, typically available within one hour. Such a tool would meet urgent clinical needs in developed countries and resource-poor settings where molecular testing is less readily available.

## Methods

### Data Collection

Linked de-identified demographic and clinical data for all patients presenting to emergency and acute medical services at Oxford University Hospitals (OUH) between 1^st^ December 2017 and 19^th^ April 2020, were extracted from EHR systems. OUH consists of 4 teaching hospitals, serving a population of 600,000 and providing tertiary referral services to the surrounding region.

For each presentation, data extracted included admission blood tests, blood gas testing, vital signs, results of SARS-CoV-2 RT-PCR assays (Public Health England designed RdRp and Abbott Architect [Abbott, Maidenhead, UK]) of nasopharyngeal swabs, and PCR for influenza and other respiratory viruses. Where available, baseline health data were included: (i) the Charlson Comorbidity index was calculated from co-morbidities recorded during all previous hospital encounters since 1st December 2017 (if any existed), and (ii) changes in blood test values relative to pre-presentation results. Patients under the age of 18, not consenting to EHR research, or who did not receive laboratory blood tests on presentation to hospital were excluded from analysis. We confined all analyses to clinical and laboratory data that are routinely available within the first hour of presentation to hospital.

Adult patients presenting prior to the 1^st^ December 2020, and therefore prior to the global outbreak, were considered as the COVID-19-negative cohort. A subset of this cohort was admitted to hospital, forming the COVID-19-negative admissions cohort. Patients presenting between the 1^st^ December and 19^th^ April 2020 with PCR-confirmed SARS-CoV-2 infection were considered the COVID-19-positive cohort, with the subset admitted considered the COVID-19-admissions cohort. Due to incomplete penetrance of testing during early stages of the pandemic and limited sensitivity of the PCR swab test, there is uncertainty in the viral status of patients presenting during the pandemic who were untested or tested negative. These patients were therefore excluded from analysis.

### Feature Sets

Five “feature sets” of clinical variables were investigated (Table 1) including presentation laboratory blood tests, point-of-care blood gas readings, changes in laboratory blood results from pre-admission baseline, vital signs and Charlson Comorbidity Index (CCI).

**Table 1:**
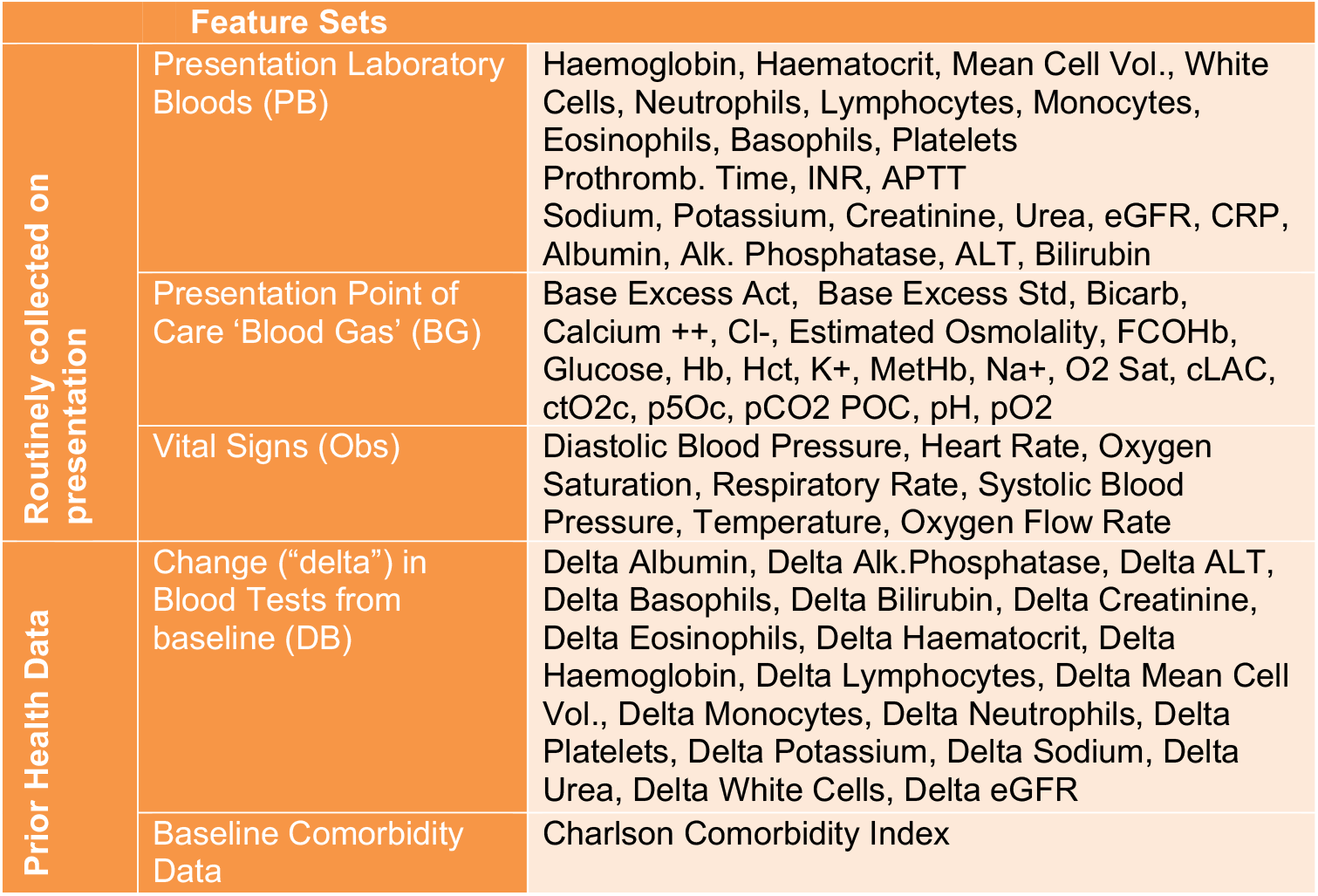
Clinical parameters included in each feature set

| Feature Sets |  |  |
| --- | --- | --- |
| Routinely collected on presentation | Presentation Laboratory Bloods (PB) | Haemoglobin, Haematocrit, Mean Cell Vol., White Cells, Neutrophils, Lymphocytes, Monocytes, Eosinophils, Basophils, Platelets<br>Prothromb. Time, INR, APTT<br>Sodium, Potassium, Creatinine, Urea, eGFR, CRP, Albumin, Alk. Phosphatase, ALT, Bilirubin |
|  | Presentation Point of Care 'Blood Gas' (BG) | Base Excess Act, Base Excess Std, Bicarb, Calcium ++, Cl-, Estimated Osmolality, FCOHb, Glucose, Hb, Hct, K+, MetHb, Na+, O2 Sat, cLAC, ctO2c, p5Oc, pCO2 POC, pH, pO2 |
|  | Vital Signs (Obs) | Diastolic Blood Pressure, Heart Rate, Oxygen Saturation, Respiratory Rate, Systolic Blood Pressure, Temperature, Oxygen Flow Rate |
| Prior Health Data | Change ("delta") in Blood Tests from baseline (DB) | Delta Albumin, Delta Alk.Phosphatase, Delta ALT, Delta Basophils, Delta Bilirubin, Delta Creatinine, Delta Eosinophils, Delta Haematocrit, Delta Haemoglobin, Delta Lymphocytes, Delta Mean Cell Vol., Delta Monocytes, Delta Neutrophils, Delta Platelets, Delta Potassium, Delta Sodium, Delta Urea, Delta White Cells, Delta eGFR |
|  | Baseline Comorbidity Data | Charlson Comorbidity Index |

Presentation blood tests and blood gas considered were result from the first blood draw on arrival to hospital, with tests not routinely available within one hour of receipt of sample excluded from analysis. Changes in blood tests were computed from pre-illness laboratory samples taken at minimum 30 days prior to presentation (available from 1^st^ December 2017 onwards). Tests where data was missing for ≥40% of all presentations were excluded and are not included in the feature sets in Table 1.

### Missing data imputation

Several imputation strategies, population mean, population median and age-based imputation, were used to impute missing data. Mean and standard deviations across imputation strategies are reported. A full description of the data processing pipeline is available in the supplementary information.

### Prediction of COVID-19 presentations

Linear (logistic regression) and non-linear ensemble (random forest & extreme gradient boosted trees, XGBoost) classifiers were trained to distinguish patients presenting or admitted to hospital with confirmed COVID-19 from pre-pandemic controls. Separate models were developed to predict COVID-19 in all patients attending the ED, and then in just the subset of those who were subsequently admitted to hospital.

### Training, Calibration and Testing

Models were trained and tested using data from 1^st^ December 2017 to 19^th^ April 2020 inclusive (Table 2). An 80:20% stratified split was performed to generate a training set and held-out test set. Using the training set, we first trained models with each independent feature set (Table 1) to identify presentations of COVID-19 from pre-pandemic controls. Next, we initialised model training using the presentation blood results feature set and sequentially added further feature sets (Table 1). Area under receiving operating characteristic curve (AUROC) achieved during training with stratified 10-fold cross validation is reported alongside standard deviations. During training, controls were matched for age, gender and ethnicity. Model thresholds were calibrated to achieve sensitivities of 70%, 80%, and 90% for identifying patients with COVID-19 in the training set prior to evaluation.

**Table 2:** Population characteristics for (a) study cohorts and (b) the independent validation set. The results are presented as percentages for categorical data and as median and interquartile range for age.

|  | (a) Study Population |  |  |  | (b) Prospective Validation Cohorts |  |
| --- | --- | --- | --- | --- | --- | --- |
|  | Presenting to Hospital |  | Admitted to Hospital |  | Presenting to Hospital | Admitted to Hospital |
| Cohort | Pre-Pandemic | COVID-19 | Pre-Pandemic | COVID-19 |  |  |
| n Patients (n COVID-19 Positive) | 114,957 (0) | 437 (437) | 71,927 (0) | 383 (383) | 3,326 (107) | 1,715 (91) |
| Age, years | 60 (38) | 69 (26) | 65 (33) | 71 (26) | 56 (37) | 64 (34) |
| Fraction male, % | 46.6 | 56.3 | 47.8 | 55.1 | 45.5 | 48.5 |
| Prior EHR Encounter, % | 74.1 | 84.0 | 74.2 | 86.4 | 80.3 | 79.7 |
| Ethnicity, % |  |  |  |  |  |  |
| White British | 76.0 | 65.4 | 78.5 | 68.4 | 66.3 | 68.2 |
| Not stated | 11.8 | 17.4 | 11.0 | 16.2 | 19.5 | 20.5 |
| Any other White background | 5.0 | 3.7 | 4.0 | 3.4 | 6.5 | 4.7 |
| Pakistani | 1.3 | 1.1 | 1.1 | 1.0 | 1.2 | 1.0 |
| Any other Asian background | 0.9 | 2.5 | 0.8 | 1.8 | 1.4 | 1.2 |
| Indian or British Indian | 0.8 | 1.1 | 0.7 | 0.8 | 0.9 | 0.8 |
| White Irish | 0.7 | 0.7 | 0.7 | 0.8 | 0.7 | 0.8 |
| African | 0.6 | 3.0 | 0.6 | 2.9 | 0.6 | 0.8 |
| Any other Black background | 0.3 | 0.9 | 0.3 | 0.5 | 0.5 | 0.3 |
| Bangladeshi | 0.2 | 0.7 | 0.2 | 0.8 | 0.3 | 0.3 |
| Chinese | 0.2 | 0.2 | 0.2 | 0.3 | 0.4 | 0.3 |
| Any other ethnic group | 2.0 | 3.2 | 1.8 | 3.2 | 1.6 | 1.3 |
| Influenza Positive | 484 | 0 | 466 | 0 | 0 | 0 |

We assessed performance of our models using the held-out test set. Firstly, we configured the test set with equal numbers of COVID-19 cases and pre-pandemic controls and reported AUROC alongside sensitivity and specificity at each calibrated threshold. Secondly, to simulate model performance at varying stages of the pandemic, we generated a series of test sets with a variety of prevalences of COVID-19 (1-50%) amongst controls using the held-out set. Positive and negative predictive values are reported for each model at the 70% and 80% sensitivity thresholds.

AUROC, sensitivity, specificity and precision are reported for candidate models at the three thresholds described above. Positive predictive value (PPV) and negative predictive value (NPV) are reported for the simulated test sets.

### Validation

Models were validated independently using data for all adult patients presenting or admitted to OUH between 20th April and 6th May 2020, by direct comparison of model prediction against SARS-CoV-2 PCR results. Due to incomplete penetrance of testing and limited sensitivity of the PCR swab test, there is uncertainty in the viral status of patients untested or testing negative. We therefore performed a sensitivity analysis to ensure disease freedom in controls, switching patients untested or testing negative with pre-pandemic ‘true-negatives’ matched for age, gender and ethnicity. Accuracy, AUROC, NPV and PPV are reported during validation.

### Ethics

The study protocol, design and data-requirements were approved by the National Health Service (NHS) Health Research Authority (IRAS ID: 281832) and sponsored by the University of Oxford.

## Data & Code availability

The data studied are available from the Infections in Oxfordshire Research Database (https://oxfordbrc.nihr.ac.uk/research-themes-overview/antimicrobial-resistance-and-modernising-microbiology/infections-in-oxfordshire-research-database-iord/), subject to an application meeting the ethical and governance requirements of the Database. Code and supplementary information for this paper are available online.

## Results

### Dataset & Cohorts

Figure 1 provides a schematic overview of cohorts in our analysis. 155,689 adult presentations were considered between 01 December 2017 and 19^th^ April 2020. 114,957 presentations to hospital prior to the 1^st^ December 2019, and therefore preceding the SARS-CoV-2 pandemic, formed the COVID-19-negative cohort. 534 patients had a RT-PCR confirmed diagnosis of COVID-19 between 1^st^ December 2019 and 19^th^ April 2020, forming the COVID-19-positive cohort. 43,378 presentations during the pandemic with no SARS-CoV-2 PCR or only negative result(s) were excluded from analysis due to uncertainty in viral status.

**Figure 1:**
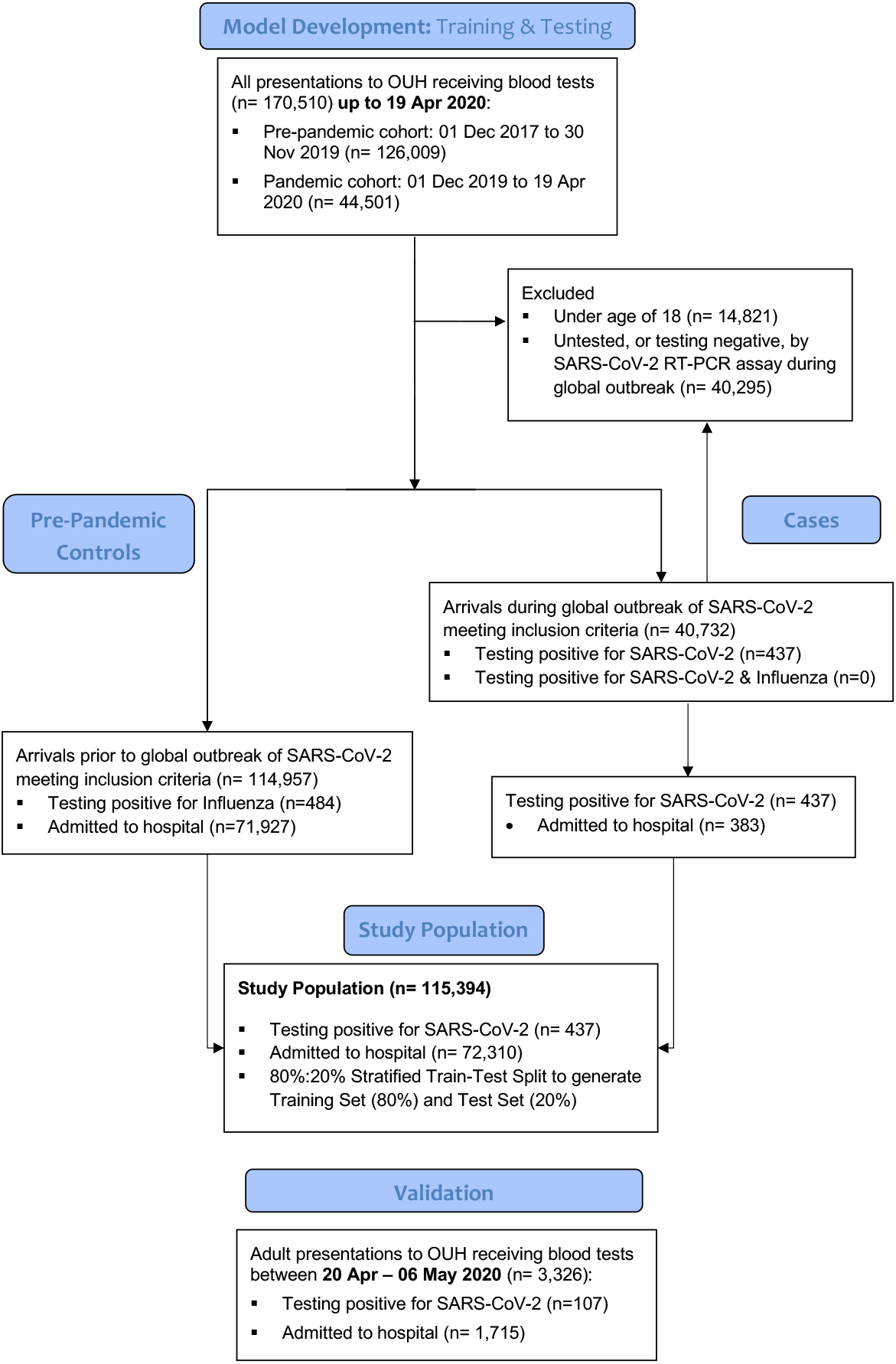
CONSORT diagram showing inclusion of patients and derivation of cohorts during model development to form (a) training and (b) test sets, and a fully independent, prospective (c) validation cohort.

Table 2 demonstrates summary characteristics of presentations included within our dataset. Patients presenting to hospital with COVID-19 had a higher median age (IQR) than pre-pandemic controls (69 (37) versus 60 (38), Kruksal-Wallis test p<0.001). Similarly, patients admitted due to COVID-19 were comparatively older, having a median age of 71 (26) versus 65 (33) for pre-pandemic admissions (p<0.001). A high proportion of patients presenting to hospital, 74.1%, had had a previous clinical encounter at the four-centre hospital group.

### Presentation bloods and vital signs are most predictive of COVID-19

Table 3 shows a summary of the relative performance of models trained using each independent feature set at identifying presentations due to COVID-19, reported in terms of AUROC achieved during stratified 10-fold cross validation alongside standard deviations (SDs). Both ensemble methods outperform logistic regression due to their intrinsic ability to detect non-linear effects of the feature sets. XGBoost classifiers trained on presentation laboratory blood tests and vital signs demonstrate highest predictive performance for COVID-19, achieving AUROCs of 0.904 (0.000) and 0.823 respectively (0.005). Narrow standard deviations demonstrate model stability.

**Table 3:**
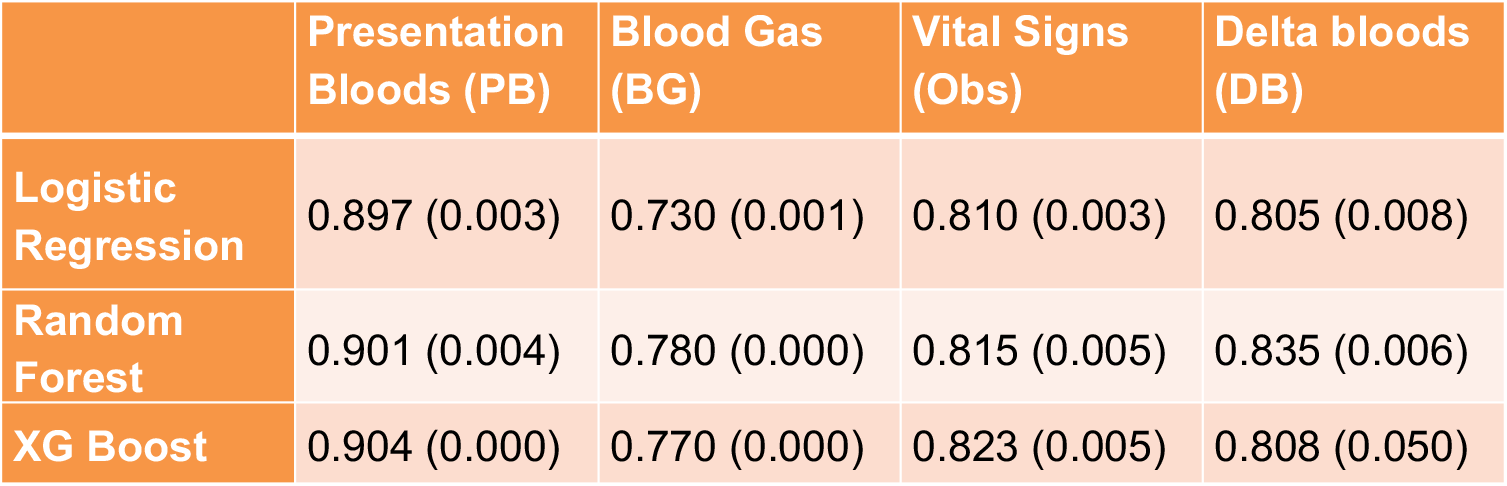
AUROC (SD) achieved for each independent feature set using stratified 10-fold cross validation during training.

### Increasing feature sets improves predictive performance for COVID-19

Stepwise addition of routinely collected clinical data supports improvement in model performance at discriminating presentations due to COVID-19 (Table 4) to a peak AUROC of 0.929 (0.003), achieved with 10-fold cross validation during training using the XGBoost classifier. Incorporating previous blood results further improves model performance to an AUROC of 0.942 (0.002), however having added previous blood tests addition of the CCI did not further improve performance.

### Developing context-specific diagnostic models

Our preliminary results (Table 4) suggest a non-linear modelling approach with clinical data routinely available on presentation (presentation blood tests, blood gas results and vital signs) achieves high classification performance (AUROC 0.929). Although incorporating prior health data supports a small increment in model performance (AUROC 0.942), missingness could limit generalisability. Detailed performance metrics for all feature set combinations, at each reported threshold, is available in the Supplementary Information.

**Table 4:**
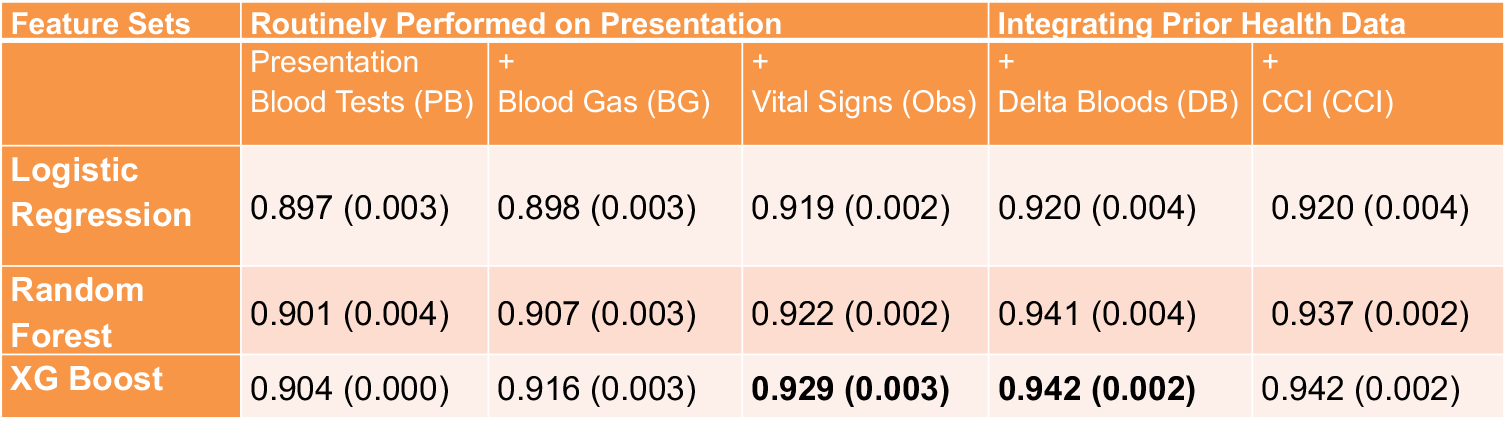
AUROC (+/- SD) achieved with increasing feature sets using stratified 10-fold cross validation during training.

We therefore developed and optimised context-specific models using the XGBoost classifier, using only clinical data sets routinely available on presentation, training separate models to predict COVID-19 in patients attending ED (ED Model) and the subset subsequently admitted to hospital (Admissions model). This approach has the advantage of requiring no previous health data, therefore being applicable to all patients, and is specific to the clinical contexts in which models use is intended.

### Our ED and Admissions models identify COVID-19 effectively in test sets of patients presenting and admitted to hospital

Performance of our ED model was assessed on a held-out test set, generated using a stratified 80%:20% test-train split of cases and configured initially with equal numbers of COVID-19 cases and pre-pandemic controls, i.e. 50% prevalence. Our ED model, calibrated during training to sensitivity of 80%, achieved an AUROC of 0.939, sensitivity of 77.4% and specificity of 95.7%.

Relative feature importance analysis demonstrated that all feature sets contributed to the most-informative variables for model predictions (Figure 3). In the ED model, three laboratory blood markers (eosinophils, basophils, and C-Reactive Protein [CRP]) were amongst the highest-ranking variables. Blood gas measurements (calcium and methaemoglobin) and vital signs (oxygen requirement and respiratory rate) were additionally amongst the variables most informative to model predictions.

**Figure 2:**
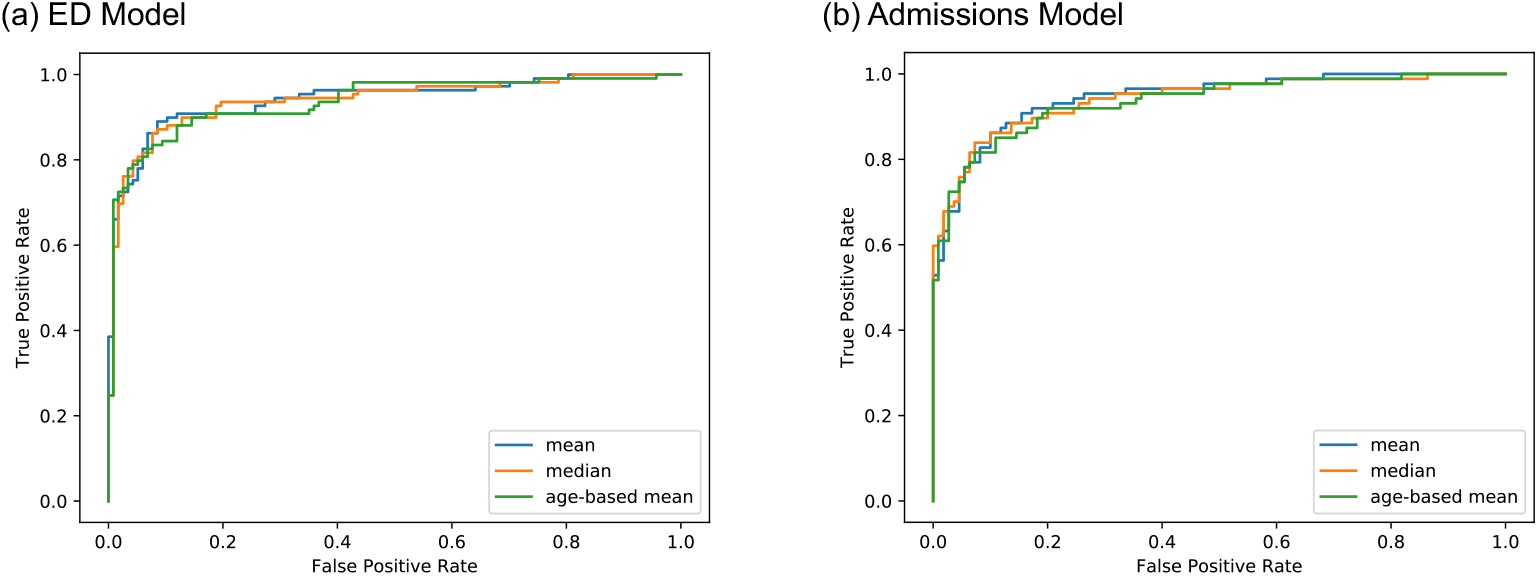
Receiver Operating Characteristic Curves for (a) our ED and (b) Admissions models.

**Figure 3:**
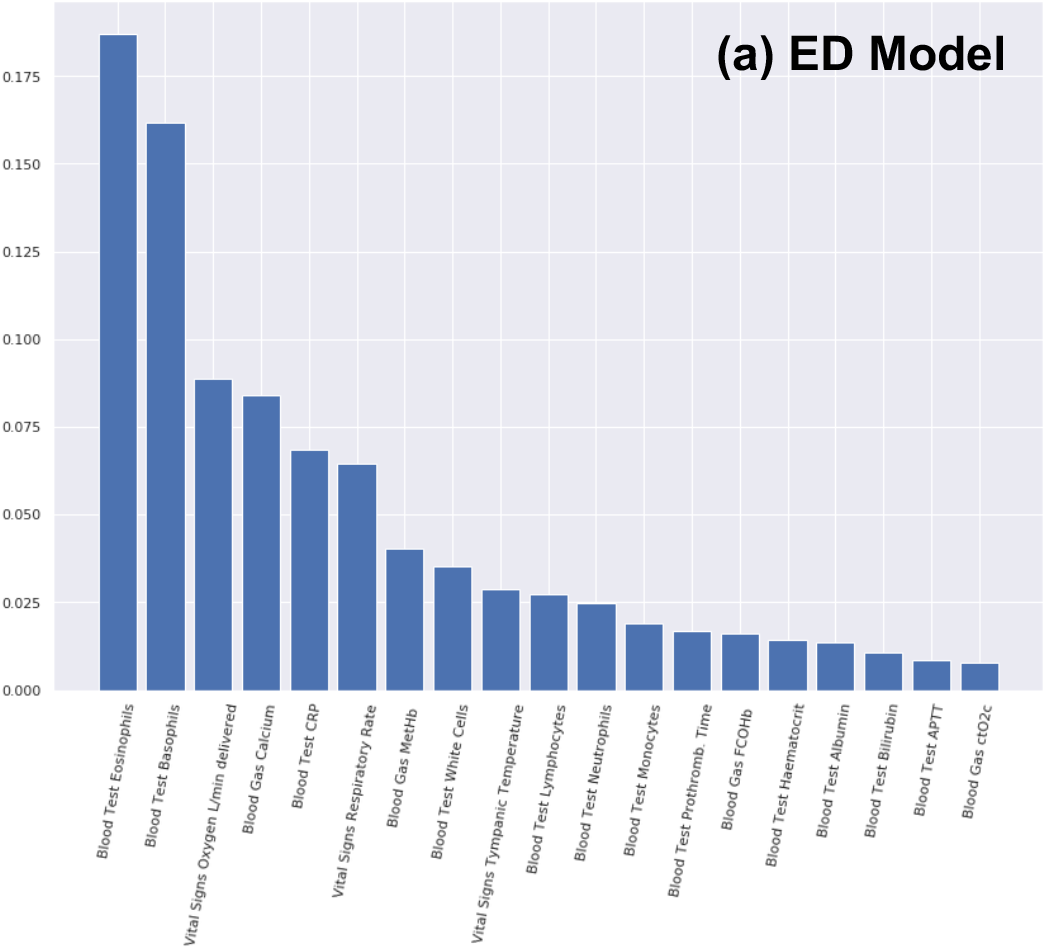
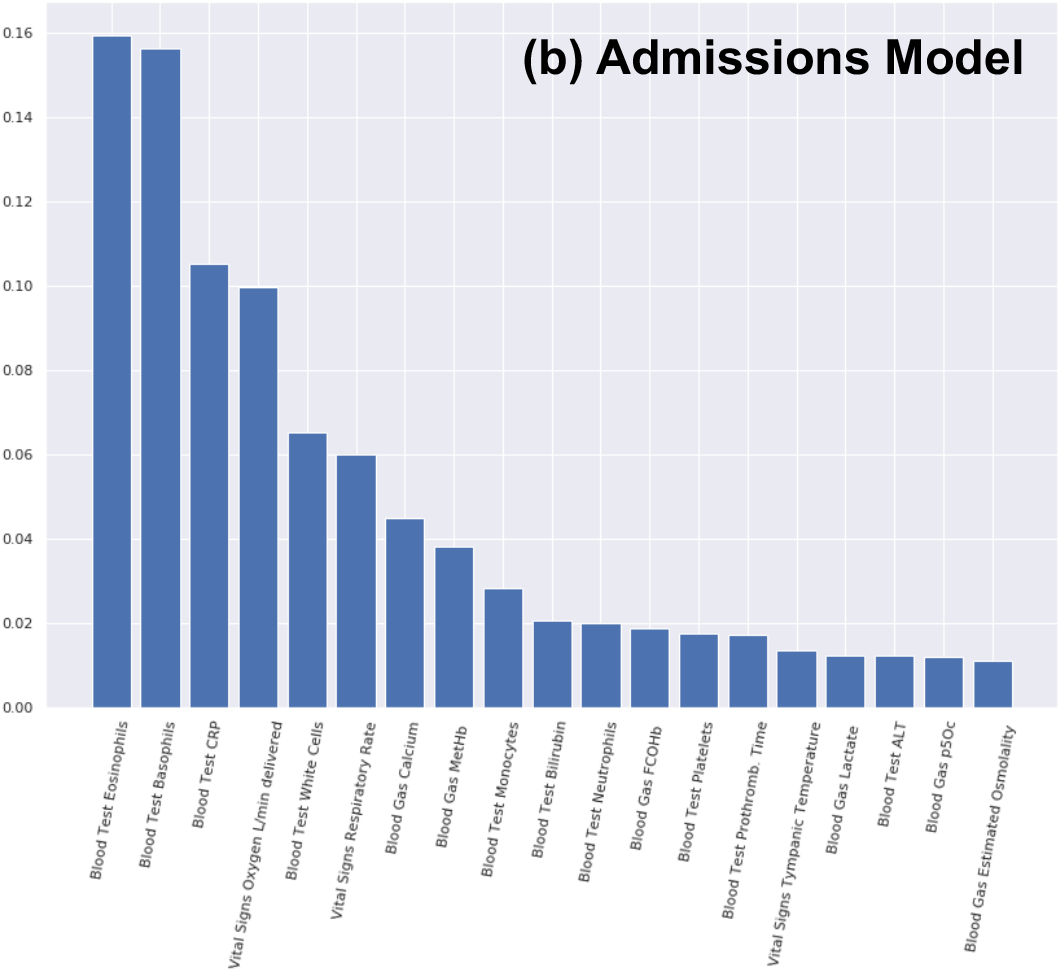
Relative feature importances within our (a) ED model and (b) Admissions model for identifying COVID-19 in patients presenting or admitted to hospital.

Similar top-ranking features are seen in the Admissions model, however notably with greater relative weights for CRP and White Cell counts and lesser weights for blood gas measurements (calcium and methaemoglobin).

### Our models achieve clinically useful predictive values at varying stages of an epidemic to support clinical decision making

To reflect performance at varying stages of an epidemic, positive and negative predictive values are assessed on test sets configured to a variety of prevalences of COVID-19. Results are reported in Table 6 for ED and Admissions models, calibrated to two sensitivity thresholds (70% and 80%).

**Table 5:**
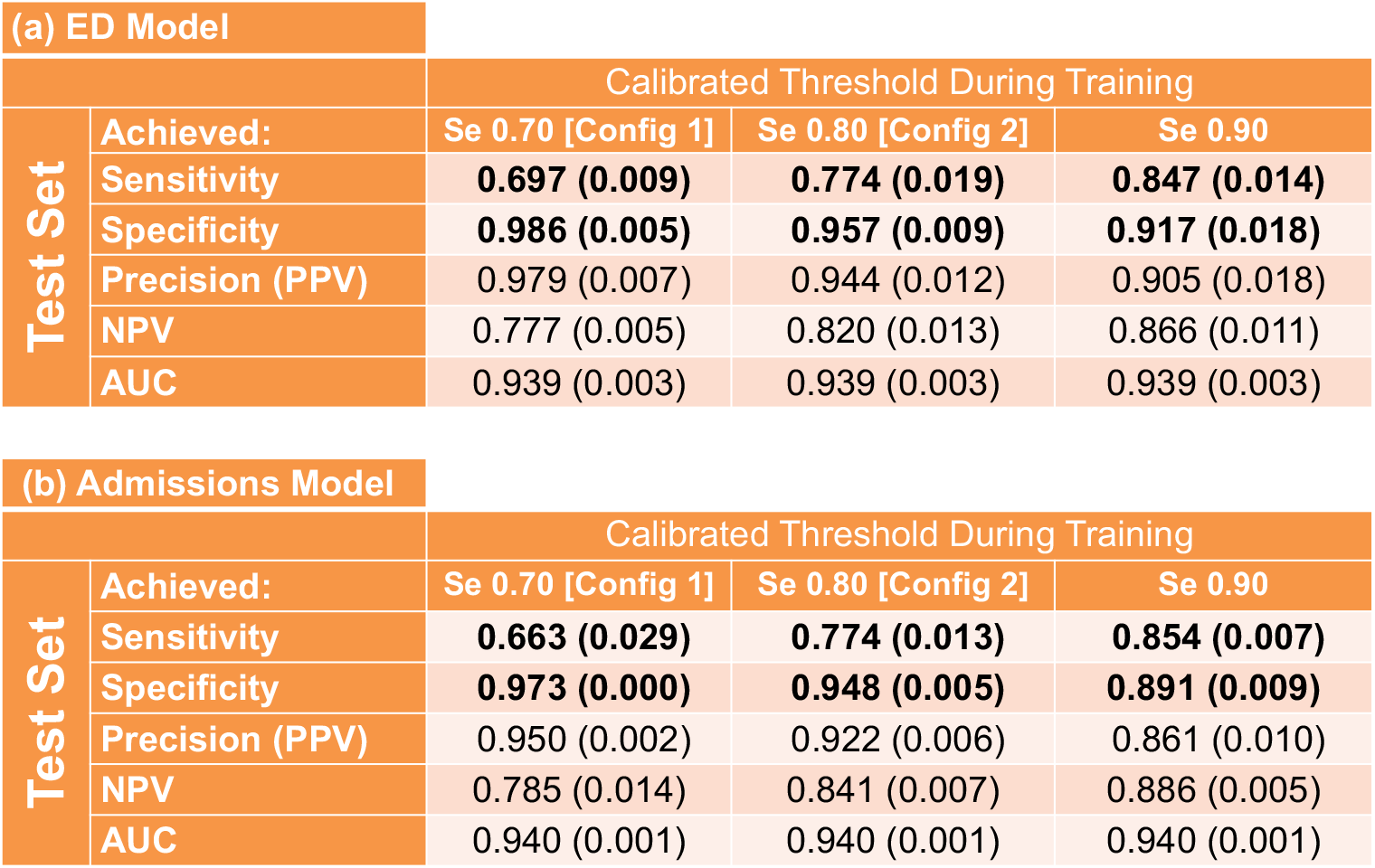
Assessment of performance (SD) of (a) our ED and (b) Admissions models, calibrated to 70, 80 and 90% sensitivities during training, at identifying COVID-19 amongst patients presenting to or admitted hospital emergency departments in a held-out test set with 50% assumed prevalence.

**Table 6:**
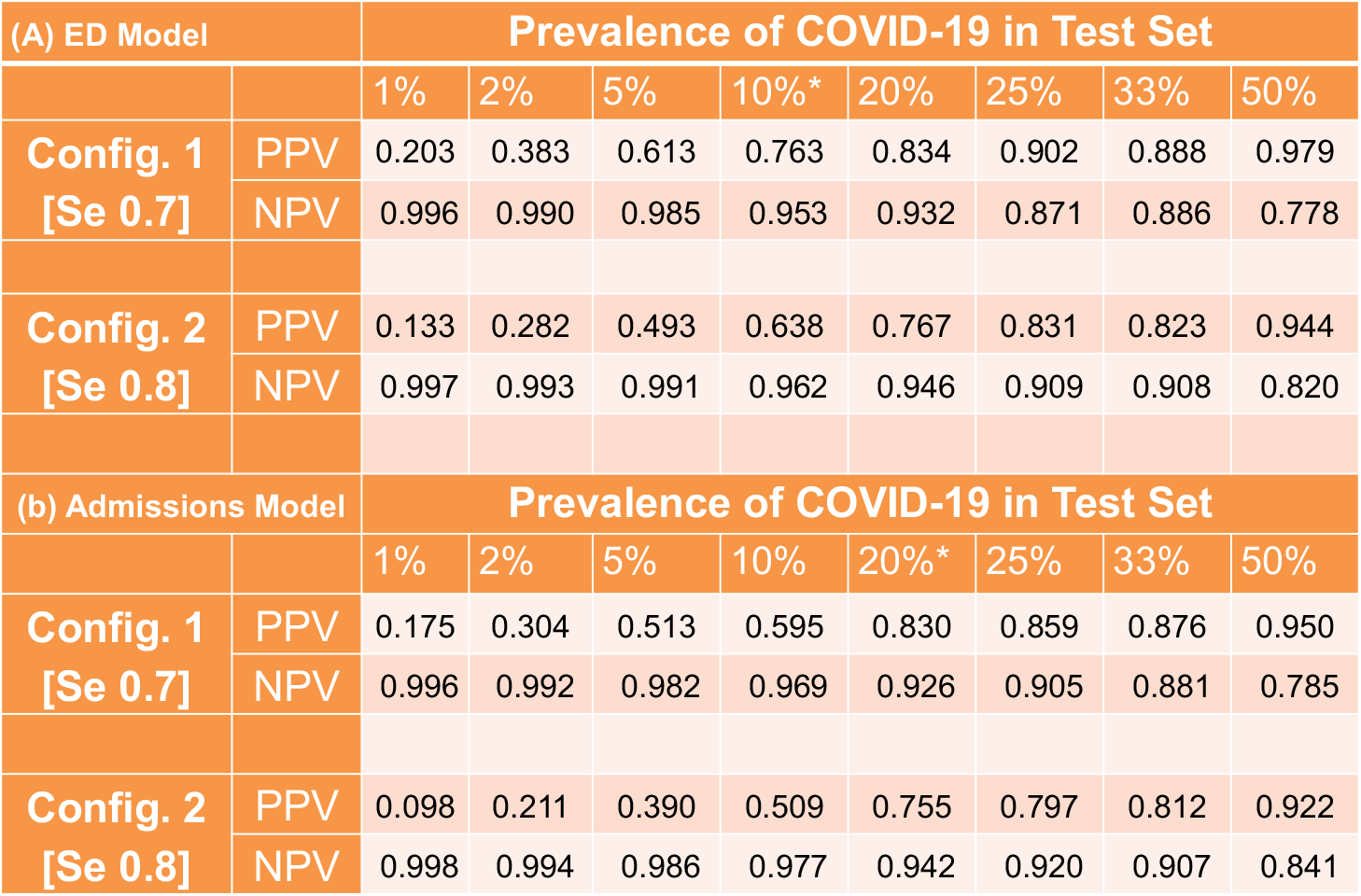
PPV and NPV of our (a) ED model and (b) Admissions model, calibrated during training to (Config. 1) 70% and (Config. 2) 80% sensitivities, for identifying COVID-19 in test sets with a variety of prevalences. The 10% and 20% scenarios (*) approximate the observed prevalence of COVID-19 in patients (a) presenting and (b) admitted to the study hospitals during the first week of April 2020 (1st April – 8th April 2020).

For both models, the higher sensitivity configuration (80%, Config 2.) achieves high NPV (>99%) where the disease is relatively uncommon (< 5% prevalence), supporting safe exclusion of the disease. At high disease prevalances (>20%), the 70% sensitivity configuration optimises for high PPV (>83%) at good NPV (>92%).

The 70% sensitivity configurations (Config. 1) of our models achieved high PPV, of 76.3% and 83.0%, and NPV, of 95.3% and 96.2%, at the prevalence of COVID-19 observed in patients presenting and admitted to hospital respectively at the study hospitals during the first week of April 2020 (1^st^ – 8th April 2020) (Table 6).

### Validation: Prospective validation of our ED & Admission models confirms high accuracy and negative predictive performance

To assess real-world performance of our ED and Admission models, calibrated during training to 80%, we validated our models for all patients presenting or admitted across the study hospital group (OUH) between 20^th^ April and 6^th^ May 2020. Prevalences of COVID-19 in patients presenting and admitted to hospital in the validation set were 3.2% and 5.3% respectively. Our ED model performed with 92.3% accuracy (AUROC: 0.881) and Admission model with 92.5% accuracy (AUROC: 0.871) on the validation set assessed against results of formal PCR testing. PPVs were 46.7% and 40.0%, and NPVs were 97.6% and 97.7% respectively.

We performed a sensitivity analysis to account for uncertainty in the viral status of patients in the validation set testing negative by PCR or who were not tested. Our ED model demonstrated an apparent improvement in accuracy to 95.1% (AUROC: 0.960), and admission model to 94.1% accuracy (AUROC: 0.937) on the adjusted validation set. NPVs achieved were also improved, at 99.0% and 98.5% respectively.

### Assessment of Misclassification

To assess for biases in model performance, we assessed rates of patient misclassification during validation of our ED and Admissions models. We observed that rates of misclassification were similar between white British (9% and 10%, respectively) and black, Asian and minority ethnic group patients (11 and 13%; Fishers’ Exact test p= 0.374 & 0.358), and between men (11% and 11%) and women (8% and 8%; p=0.147 and 0.091). We also found no difference between misclassification of patients aged over 60 (10% and 10%) and patients aged between 18 and 60 (9% and 8%; p=0.187 & 0.191).

## Discussion

Limitations of the gold-standard PCR test for COVID-19 have challenged healthcare systems across the world. There remains an urgent clinical need for rapid and accurate testing on arrival to hospitals, with the current test limited by prolonged turnaround times^18^, shortages of specialist equipment and operators, and relatively low sensitivity^8^.

In this study, we develop and assess two Artificial Intelligence (AI) driven screening tools for in-hospital COVID-19 screening, in the clinical context intended for use. Our Emergency Department and Admission models effectively identify patients with COVID-19 amongst all patients presenting and admitted to hospital, using data typically available within the first hour of presentation (AUROC 0.939 & 0.940). On validation using appropriate prospective cohorts of all patients presenting or admitted to a large UK teaching centre group between 20^th^ April and 6^th^ May 2020 (n=3,326 & 1,715), our models achieve high accuracies (92.3% and 92.5%) and negative predictive values (97.6% and 97.7%). A sensitivity analysis to account for uncertainty in negative PCR results improves apparent accuracy (95.1% and 94.1%) and NPV (99.0% and 98.5%). Simulation on test-sets with varying prevalences of COVID-19 shows that our models achieve clinically useful negative predictive values (>0.99) at low prevalences (<5%), supporting safe exclusion of the disease. At higher prevalences (>25%), our models can be configured to meet clinical needs for higher positive predictive values (>0.83). Our models’ negative predictive performance supports use as a screening test to rapidly exclude COVID-19 in emergency departments, assisting immediate care decisions, guiding safe patient streaming and serving as a pre-test for formal RT-PCR testing where availability is limited.

Strengths of our AI approach include an ability to scale rapidly to meet the urgent clinical need, taking advantage of cloud computing platforms, and working with laboratory tests widely available and routinely performed within the current standard of care. Moreover, at higher prevalances of COVID-19, clinical need may favour higher PPV; we demonstrate that our models can be calibrated to meet changing clinical requirements as the pandemic progresses.

To date early-detection models have overwhelmingly focussed on assessment of radiological imaging, such as Computerised Tomography (CT)^5,18–20^, that is less readily available and involves patient exposure to ionising radiation. Few studies have assessed routine laboratory tests, with studies to-date including small numbers of confirmed COVID-19 patients, using RT-PCR results for data labelling thereby failing to ensure disease freedom in ‘negative’ patients, and are not validated in the clinical population intended for use^11,12,21^. A significant limitation of existing works is the use of narrow control cohorts during training, inadequately exposing models to the breadth and variety of alternative infectious and non-infectious pathologies, including seasonal pathologies. Moreover, though the use of AI techniques for early detection holds great promise, many published models to date have been assessed to be at high risk of bias^19^.

Our study includes the largest dataset of any COVID-19 laboratory AI study to date, considering over 115,000 hospital attendances and 5 million laboratory measurements, and is prospectively validated using the appropriate patient cohorts for the models’ intended clinical contexts. The breadth of our pre-pandemic control cohort gives exposure to a wide range of undifferentiated presentations, including other seasonal infectious pathologies (e.g. Influenza), and offers confidence in SARS-CoV-2 freedom. Additionally, our study is the first to integrate presentation laboratory blood results with blood gas and vital signs measurements, maximising richness of the dataset available within the acute clinical setting.

Our results demonstrate that integrating prior health data, such as calculated differences in blood tests, incrementally improved performance of our ED and Admission models (AUROC 0.944 and 0.946, Supplementary Information). However, prior health data was unavailable for 15.6% of patients presenting with COVID-19 (Table 2), and may be less readily available at other sites. As clinically adequate performance was achieved on presentation data alone, without compromising generalisability, we did not include prior health data in our final models.

We select interpretable linear and non-linear modelling approaches, achieving highest performance with extreme gradient boosted tree methods. Information variables from all sets were important in model predictions, including three measured biochemical quantities (Eosinophils, Basophils and CRP), blood gas measurements (Methaemoglobin and Calcium), and vital signs (Respiratory Rate and Oxygen Delivery). Where features are highly correlated, any one of the correlated features may be selected during training and ascribed importance. After selecting one such feature, the relative importance of other correlated features is decreased as the relationship is encoded within the value of the selected correlate. Interpretation of significance for variable absence should therefore be cautious.

Existing literature has reported an association between lymphopenia and COVID-19 ^3,17^. We observe that lymphopenia is frequently absent on first-available laboratory tests performed on admission (Supplementary Information, Table C1), and is not a highly-ranked feature in our models (Figure 3). Univariate analysis identifies that low Eosinophil count on presentation is more strongly correlated with COVID-19 diagnosis than the Lymphocyte count (Supplementary Information, Appendix B; chi-squared scores 41.61 and 31.56 respectively).

Recognising concerns of biases within AI models, we assessed cases misclassified during validation for evidence of ethnic, age or gender biases. Our results showed misclassification was equally likely between white British and black, Asian and minority ethnic patients, males and females and elderly (>60) and younger (18-59) patients.

Our study seeks to address limitations common to EHR research. We use multiple imputation for missing data, taking a mean of three strategies (age-based imputation, population mean, population median). We queried whether our results were sensitive to imputation strategy and found similar model performance across the three strategies.

A potential limitation of the present study is the relatively limited ethnic diversity of patients included. 76.0% of patients presenting to the hospital group prior to the pandemic, and 65.4% of patients with confirmed COVID-19, reported their ethnicity to be white British (Table 2). Although our models do not appear to be more likely to misclassify ethnic minority patients, integrating data from international centres would increase confidence in model generalisability.

Our work demonstrates that an AI-driven screening test can effectively triage patients presenting to hospital for COVID-19 while confirmatory laboratory PCR testing is awaited. Our approach is rapidly scalable, fitting within the existing laboratory testing infrastructure and standard of care, and additionally serves as proof-of-concept for a rapidly deployable software tool in future pandemics.

Prospective clinical trials would further assess model generalisability and real-world performance.

## Data Availability

The data studied are available from the Infections in Oxfordshire Research Database, subject to an application meeting the ethical and governance requirements of the Database.

https://oxfordbrc.nihr.ac.uk/research-themes-overview/antimicrobial-resistance-and-modernising-microbiology/infections-in-oxfordshire-research-database-iord/

## Abbreviations

AI: Artificial Intelligence
AUROC: Area under receiver operating characteristic curve
COVID-19: Coronavirus Disease 2019
CCI: Charlson Comorbidity Index
CRP: C-Reactive Protein
EHR: Electronic Health Records
LR: Logistic Regression
NPV: Negative Predictive Value
OUH: Oxford University Hospitals NHS Foundation Trust
POCT: Point of Care Test
PPV: Positive Predictive Value
RF: Random Forest
RT-PCR: Real Time Polymerase Chain Reaction
SARS-CoV-2: Severe Acute Respiratory Syndrome Coronavirus 2

## Acknowledgments

We express our sincere thanks to all patients, clinicians and support staff across Oxford University Hospitals NHS Foundation Trust. We additionally thank staff across the University of Oxford Institute of Biomedical Engineering, Research Services and Clinical Trials & Research Group. In particular, we thank Corinne Prescot for her assistance administering the study, Dr Ravi Pattanshetty for clinical input, and Jia Wei.

## Declarations

DWE reports lecture fees from Gilead, outside the submitted work. DC reports Consultancy for Oxford University Innovation, Biobeats, and Sensyne Health. No other authors report any conflicts of interest.

## Contributions

AS, DC, TZ, DE, ZBH, TP conceived of and designed the study. DWE extracted the data from EHR systems. TT, SK, AS pre-processed the data. DK, SK, AS, TZ, TT, DWE, DC developed the models. AS, SK, DK, TZ, AB, DWE, DC validated the models. AS, SK, ZBH wrote the manuscript. All authors revised the manuscript.

## Supplementary Material

## Appendix A Additional Methods & Data Processing

### Data Extraction

De-identified demographic, microbiology and laboratory records from the first 24 hours of presentation to the hospital were extracted retrospectively from electronic health records for all cohorts. Where available, pre-morbid blood tests were extracted for patients, dated at minimum 30 days prior to acute presentation to hospital. The curated data set included 71 features (24 laboratory blood tests, 21 Blood gas readings, six routinely measured physiological parameters, 19 changes in laboratory blood results from the baseline, and the Charlson Comborbidity Index), for 170,510 hospital presentations across a four-site NHS trust in Oxfordshire, UK.

### Data Cleaning

Non-numerical readings were replaced with clinically appropriate values. Where a lab value was reported as being below the threshold of detection of the laboratory assay, the value was replaced with a numerical zero value. Where values were reported as being above the threshold of detection, clinically appropriate values were selected to maintain the significance of the high result. The distribution of features in terms of mean and interquartile ranges can be seen in Tables C1-C3.

**Table C1.**
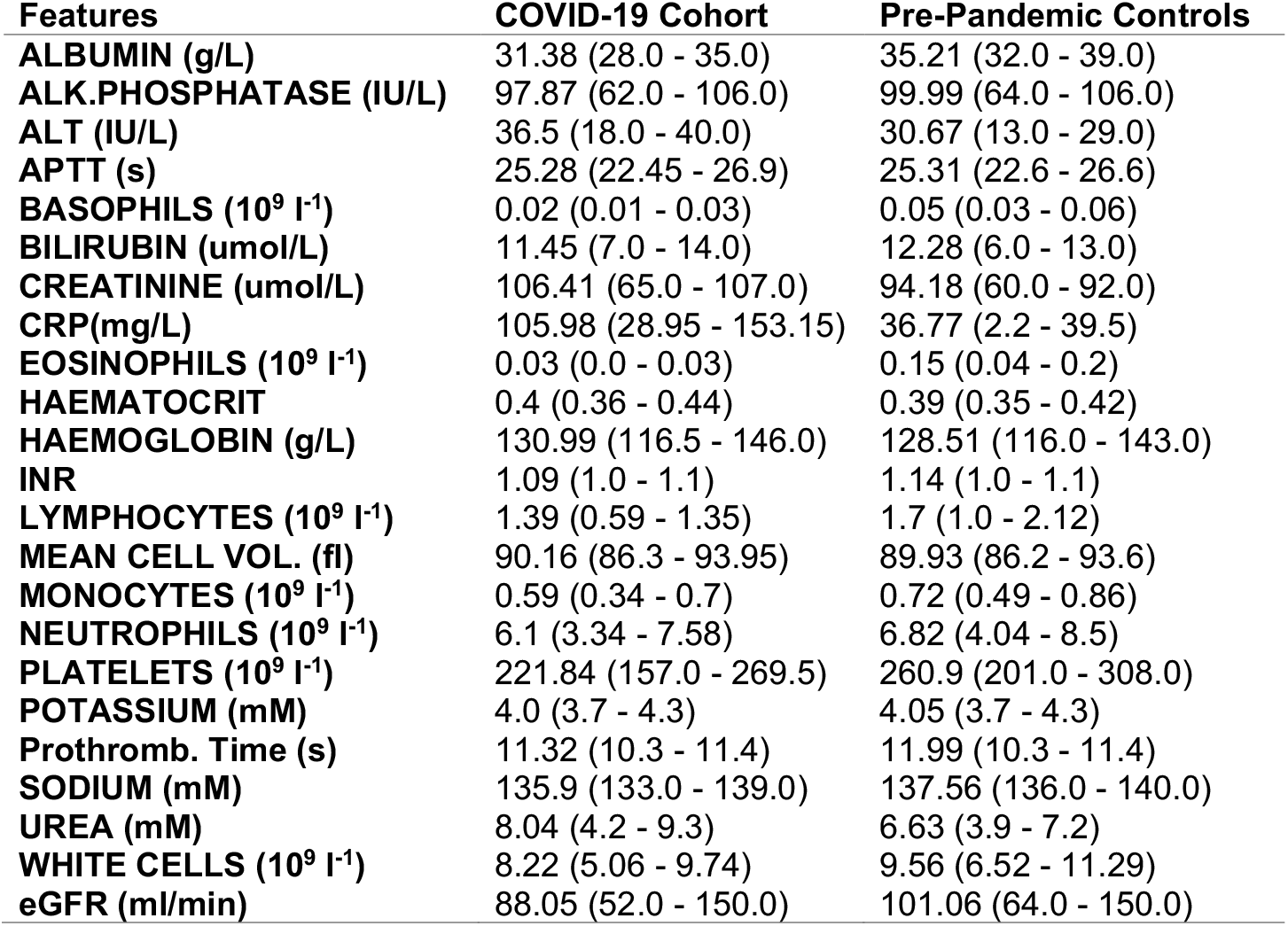
Distribution of the Blood Test features reported as mean and interquartile ranges for COVID-19 cases and pre-pandemic controls

| Features | COVID-19 Cohort | Pre-Pandemic Controls |
| --- | --- | --- |
| ALBUMIN (g/L) | 31.38 (28.0 - 35.0) | 35.21 (32.0 - 39.0) |
| ALK.PHOSPHATASE (IU/L) | 97.87 (62.0 - 106.0) | 99.99 (64.0 - 106.0) |
| ALT (IU/L) | 36.5 (18.0 - 40.0) | 30.67 (13.0 - 29.0) |
| APTT (s) | 25.28 (22.45 - 26.9) | 25.31 (22.6 - 26.6) |
| BASOPHILS ($10^9 \text{ l}^{-1}$) | 0.02 (0.01 - 0.03) | 0.05 (0.03 - 0.06) |
| BILIRUBIN ($\mu\text{mol/L}$) | 11.45 (7.0 - 14.0) | 12.28 (6.0 - 13.0) |
| CREATININE ($\mu\text{mol/L}$) | 106.41 (65.0 - 107.0) | 94.18 (60.0 - 92.0) |
| CRP(mg/L) | 105.98 (28.95 - 153.15) | 36.77 (2.2 - 39.5) |
| EOSINOPHILS ($10^9 \text{ l}^{-1}$) | 0.03 (0.0 - 0.03) | 0.15 (0.04 - 0.2) |
| HAEMATOCRIT | 0.4 (0.36 - 0.44) | 0.39 (0.35 - 0.42) |
| HAEMOGLOBIN (g/L) | 130.99 (116.5 - 146.0) | 128.51 (116.0 - 143.0) |
| INR | 1.09 (1.0 - 1.1) | 1.14 (1.0 - 1.1) |
| LYMPHOCYTES ($10^9 \text{ l}^{-1}$) | 1.39 (0.59 - 1.35) | 1.7 (1.0 - 2.12) |
| MEAN CELL VOL. (fl) | 90.16 (86.3 - 93.95) | 89.93 (86.2 - 93.6) |
| MONOCYTES ($10^9 \text{ l}^{-1}$) | 0.59 (0.34 - 0.7) | 0.72 (0.49 - 0.86) |
| NEUTROPHILS ($10^9 \text{ l}^{-1}$) | 6.1 (3.34 - 7.58) | 6.82 (4.04 - 8.5) |
| PLATELETS ($10^9 \text{ l}^{-1}$) | 221.84 (157.0 - 269.5) | 260.9 (201.0 - 308.0) |
| POTASSIUM (mM) | 4.0 (3.7 - 4.3) | 4.05 (3.7 - 4.3) |
| Prothromb. Time (s) | 11.32 (10.3 - 11.4) | 11.99 (10.3 - 11.4) |
| SODIUM (mM) | 135.9 (133.0 - 139.0) | 137.56 (136.0 - 140.0) |
| UREA (mM) | 8.04 (4.2 - 9.3) | 6.63 (3.9 - 7.2) |
| WHITE CELLS ($10^9 \text{ l}^{-1}$) | 8.22 (5.06 - 9.74) | 9.56 (6.52 - 11.29) |
| eGFR (ml/min) | 88.05 (52.0 - 150.0) | 101.06 (64.0 - 150.0) |

**Table C2.** Distribution of the Blood Gas features reported as mean and interquartile ranges for COVID-19 cases and pre-pandemic controls

| Features | COVID-19 Cohort | Pre-Pandemic Controls |
| --- | --- | --- |
| BE Act (mM) | 0.97 (-0.65 - 3.0) | 0.66 (-0.8 - 2.5) |
| BE Std (mM) | 1.1 (-0.8 - 3.3) | 1.03 (-0.8 - 3.2) |
| BICARB (mM) | 24.64 (23.2 - 26.3) | 24.46 (23.2 - 26.0) |
| Ca+ + (mM) | 1.12 (1.08 - 1.16) | 1.18 (1.14 - 1.22) |
| Cl- (mM) | 102.41 (99.0 - 106.0) | 104.21 (102.0 - 107.0) |
| Estimated Osmolality | 281.08 (274.25 - 287.25) | 283.72 (278.7 - 289.4) |
| FCOHb (%) | 0.87 (0.6 - 1.1) | 1.32 (0.7 - 1.4) |
| Glucose (mM) | 8.11 (5.7 - 8.6) | 7.15 (5.4 - 7.5) |
| Hb (g/L) | 136.4 (124.0 - 152.0) | 134.62 (122.0 - 149.0) |
| Hct | 41.86 (38.0 - 46.5) | 41.26 (37.5 - 45.7) |
| K+ (mM) | 3.95 (3.6 - 4.2) | 4.06 (3.7 - 4.3) |
| MetHb (%) | 0.62 (0.4 - 0.8) | 0.88 (0.6 - 1.1) |
| Na+ (mM) | 136.11 (133.0 - 140.0) | 137.82 (136.0 - 141.0) |
| O2 Sat (%) | 60.18 (37.95 - 83.15) | 63.25 (43.8 - 84.3) |
| cLAC (mM) | 1.8 (1.1 - 2.0) | 1.65 (0.9 - 1.9) |
| ctO2c | 11.4 (7.1 - 15.3) | 11.71 (7.9 - 15.5) |
| p5Oc (kPa) | 3.73 (3.5 - 3.95) | 3.73 (3.5 - 3.93) |
| pCO2 (kPa) | 5.35 (4.59 - 5.97) | 5.64 (4.95 - 6.25) |
| pH | 7.42 (7.38 - 7.46) | 7.4 (7.37 - 7.43) |
| pO2 (kPa) | 5.25 (3.31 - 6.36) | 6.13 (3.49 - 6.62) |

**Table C3.**
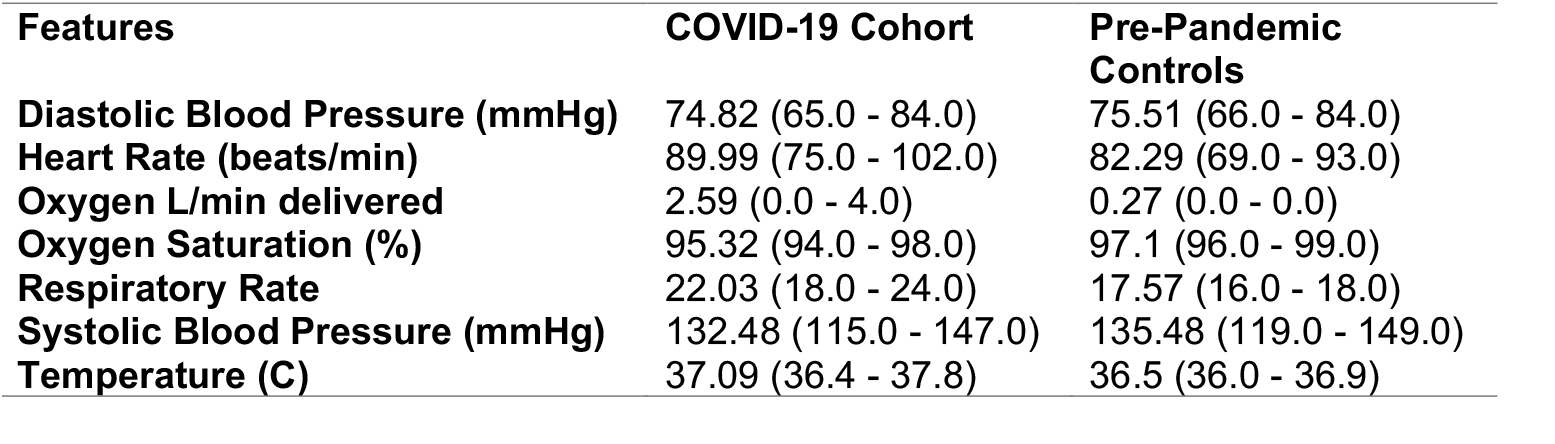
Distribution of the Vital Sign features in terms of mean and interquartile ranges for positive and control cases

### Missing Data

Multiple imputation strategies, population mean, population median and age-based imputation, were used to impute missing data. The data was analysed by all three methods individually and the mean performance was reported.

### Normalisation

Data normalisation was implemented to mitigate overfitting and to avoid the reliance of the model on measurement units. Categorical data are handled by encoding as “1-hot” variables.

### Methodology

Three machine learning techniques, logistic regression (LR), random forest (RF), and Gradient Boosting Tree (XGB), were considered and compared in terms of predictive performance. LR is a linear model that optimises a set of weights for each feature to achieve the best classification performance on the training data. This model was built using the LIBLINEAR library. LR is easy to implement, efficient to train and provides probabilities as outcomes. However, LR has high bias and cannot solve non-linear problems due to its linear decision surface. RF is an averaging method that is based on building several independent classifiers. This model fits several decision tree (DT) classifiers on different subsets of the dataset and averages results to produce final predictions with improved performance. RF is based on training several independent trees that can be fit in parallel, and often reduces the variance, however, can be more computationally expensive. 100 estimators were considered for RF training. XGBoost is a generalisation of boosting to an arbitrary differentiable loss function. XGBoost is more robust to outliers and has high predictive power. Nonetheless, due to its sequential nature, it cannot be parallelised. DT was used as the base classifier and Binomial deviance and 100 estimators were considered for the training.

## Appendix B Univariate Chi Sqaured test

Statistical tests such as chi Square can be used to select features that have a strong correlation with the outcome. The chi-squared (chi^2^) is a statistical test for non-negative features. The importance based on chi^2^ for three important blood test markers (based on our results and the literature) can be seen in Table C4.

| Feature | Score |
| --- | --- |
| EOSINOPHILS | 47.61 |
| LYMPHOCYTE | 31.56 |
| BASOPHILS | 5.92 |

## Appendix C Detailed Results

The full results for various feature sets are attached in the ‘Initial Experiments’, ‘Emergency Department Model’ and ‘Admission Model’ directories. Patient data collected between 1st December 2017 and 19th April 2020 was separated in to training and test sets by 80%:20% split. The performances were reported in terms of accuracy, area under the roc curve (AUC), precision (or PPV), recall, specificity, F1-score, and NPV. The mean and standard deviation (SD) on the held-out test sets for various prevalences and thresholds (the threshold was set to have a fixed sensitivity on the train set, e.g., 80% and was used for the hold-out test set) were reported.

Validation was performed using all patients presenting or admitted to OUH between 20th April and 6th May 2020 (‘Validation’ subfolder). Due to incomplete penetrance of testing for COVID-19 and the limited sensitivity of the RT-PCR swab test, there is uncertainty in the viral status of patients untested, or testing negative, in the prospective cohort. We therefore switched patients testing negative, or not tested, for COVID-19 in the test set with unseen, matched pre-pandemic controls (matched for age, gender and ethnicity) to ensure disease freedom (‘Adjusted Validation’ subfolder).

Furthermore, relative feature importances are reported for each experiment. The feature ranking is based on the importance of each feature in construction of the DTs within the model.

